# Mental health clinicians’ information-seeking and use of Wikimedia platforms

**DOI:** 10.1101/2024.09.17.24313842

**Authors:** Sophia Young, Eric A. Youngstrom, Anna Van Meter, Andrea S. Young, Martha A. Turner-Quest, Megan Brady, Emily M. Becker-Haimes

## Abstract

Internet sites are increasingly being used to disseminate clinical resources to mental health clinicians. Wikimedia platforms (e.g., Wikipedia and Wikiversity) are open-source, freely available resources that are among the most visited websites and could be an effective way to disseminate mental health information. An important first step in learning how to optimize the impact of such dissemination efforts is to understand how and for what reasons practicing clinicians already engage with these resources. Using a convenience sample of 120 practicing mental health clinicians (82.5% female, 85.6% white, *M* age = 41.31), we assessed clinician-reported practices about where and how they seek information about psychological science on the internet. Our results showed that freely available resources related to mental health are not a primary source of information for mental health clinicians and that clinicians have low confidence in the veracity of information available through Wikipedia and Wikiversity. Clinicians shared strategies (e.g., implementing verification steps on Wiki platform pages) that could increase clinician confidence in the information provided and their likelihood of using these sites as resources. Overall, this study indicates that mental health clinicians are not regularly using Wiki sites for mental health-related questions, even though they are accessible, regularly updated, and increasingly reliable. Findings suggest that implementing verification steps on Wiki pages could increase clinicians’ confidence in the information provided and their likelihood in using these sites as resources, however, it remains unclear whether verification steps would lead to more frequent use or how such a system would be implemented through existing platforms. Results will help guide future dissemination efforts to increase the availability and utilization of evidence-based psychological science online for clinicians.

**Author Summary:** Over the past few decades, we have seen a boom in psychological research and innovation. However, the results of this work exist primarily behind academic firewalls, making it difficult for clinicians to access information describing advancements in their field. As clinicians are best positioned to immediately address the global mental health crisis, their inability to make use of the most up-to-date research has a significant, negative impact on the quality of available mental healthcare. Wikipedia and Wikiversity present a possible solution: they are among the most utilized information sites in the world and could be a great tool for providing mental health clinicians with evidence-based psychological resources. In this study, we surveyed clinicians about their online information-seeking habits and found that Wiki platforms are not a main source of information for clinicians, who reported low confidence in the quality of information on these sites. However, survey results also indicated that implementing verification steps on Wiki pages could increase clinicians’ confidence and willingness to use them. Our results provide guidance on how evidence-based psychological science can be made available on the internet in a way that will increase its utilization and best support clinicians.

---

Research productivity in psychology and behavioral health has expanded dramatically in recent years [1]. A search of ProQuest’s database shows 12,470 results of articles with the keyword “anxiety treatment” (search narrowed to scholarly journals only) during the year 2010. PubMed’s database for 2010 has 5,131 results for “anxiety treatment”. In contrast, 13 years later, ProQuest’s database for 2023 alone shows 46,304 results for the same keyword, while PubMed yields 13,150. Similar increases exist across both websites for related topics, such as “psychotherapy,” “cognitive behavioral therapy,” and “depression treatment.” In many ways, this suggests that the field is evolving, growing, and becoming more rigorous. At the same time, research incentives can be such that people publish work of questionable quality, leaving fewer studies well-designed and clinically relevant [2], requiring consumers of research literature to sift through many studies to discern the latest scientific recommendations for mental health care. The fact that *volume* is not the same thing as *quality* increases the burden of information overload, requiring more effort to find helpful and actionable knowledge.

While research productivity has grown, so too has the demand for mental health professionals, a majority of whom work in community settings. Most of the innovations in clinical science are published in academic journals that often exist behind firewalls making them inaccessible to clinicians working outside academic institutions (who constitute the largest segment of mental health professionals). Making use of up-to-date clinical research recommendations is especially difficult for those in front-line clinical positions, who have limited time to identify and integrate information about recommended practices [3,4].

Unlike the academic psychotherapy literature, much clinical content is widely available and accessible on the internet (e.g., through search engines): 95% of U.S. adults reported using the internet in some capacity in 2023 [5]. Search results also include generally easy-to-read information. However, many of these resources may be out-of-date, as it can take a long time for new work to become public knowledge [6,7]. Large portions of the search results are driven by commercial interests, which frequently disguise their agenda, and other reputable-looking sources may not clearly disclose their advertising or financial relationships that may influence their content [8]. Also included in the mix are blogs and other sites of variable quality that would, as a group, score low on ratings for quality of research support or clinical evidence [2].

In contrast, open-source platforms like Wikipedia and Wikiversity invite frequent updates and have the capability to maintain pace with academic work [9]. Wikipedia functions as a free online encyclopedia with articles continuously edited by volunteers across the world, aiming to update pages with reliable information by verifiable sources [10]. Wikiversity is similarly edited but revolves around providing materials for continued learning, rather than functioning as an online encyclopedia [11]. Due to their dissemination processes, Wikipedia and Wikiversity can be more up to date than traditional academic journals. Though many other open-source platforms exist that target academic audiences in particular (e.g. Open Science Framework, arXiv, BioMed Central), Wikimedia sites are more widely used. Wikipedia remains one of the most consulted health resources in the world [10,12–14]. It is the seventh most popular website globally (with no other text-based information source website ahead of it), receiving between 4 to nearly 5 billion unique worldwide visitors every month [15,16]. Furthermore, due to a special relationship with Google [17], Wikipedia pages are frequently one of the top five results in internet searches [10]; as a result, Wikipedia’s English-language medical content has more traffic than WebMD and Mayo Clinic [10].

Not only do these platforms receive a lot of views, there also is some evidence that information provided on Wikipedia can directly influence human behavior. Recent studies have shown that enhancing information on certain cities’ Wikipedia pages leads to increases in tourist visits [18], and that language used in scientific Wikipedia articles impact language used in future published scientific works [19].

As the impact and use of Wikimedia platforms increases, some professional groups have consciously switched to focusing on Wikipedia pages as a way of disseminating and updating information, with epidemiology providing a leading example. The World Health Organization (WHO) partnered with the Wikimedia Foundation in 2020 to share WHO infographics and other public health information about COVID-19 on Wikipedia, making sure pandemic-related information was reliable and up to date [20]. There also has been a concerted effort and substantial push across academic and medical disciplines for more professionals and students to edit information on Wikipedia articles in events such as edit-a-thons and as course assignments in health professional schools [10,20–24]. This allows students and professionals to apply their knowledge while increasing access to accurate information for medical students, providers, and laypeople [25–27].

However, due to its open-source nature and emphasis on collaboration, the quality of the information on Wikipedia has been a source of skepticism for many professionals and academics. Concerns include reliability, potential connections to plagiarism, lack of consultation with field experts, and perceptions that it is not rigorous [21,28,29]. This stigma remains despite research suggesting that the quality of information matches or exceeds the scope and accuracy of Encyclopedia Britannica in multiple independent evaluations [30]. Furthermore, information about mental health disorders on Wikipedia is comparable to or better than what is provided by centrally controlled websites and textbooks [31] and the accuracy of Wikipedia for topics related to medicine is close to or on par with professional sources [10,32]. In politics, current events, and other areas, Wikipedia has become recognized as one of the more consistently balanced venues, and “neutral point of view” is a central tenet [8]. However, valid concerns remain about readability (i.e., how decipherable the pages are), which is an ongoing issue with Wikipedia [10,31,33] and could decrease user engagement or opinions about the content provided.

The stigma surrounding open-source platforms may further be decreasing post-pandemic in the wake of increasing desire for transparent science [34] alongside efforts to legitimize the information provided on open-source platforms [25,28,29,25]. The Wikimedia Foundation has also developed strict quality control processes [10,21] and has created the WikiJournal of Science, a peer-reviewed open-access journal covering topics in science, technology, engineering, and mathematics that is rapidly integrated into corresponding Wikipedia pages once published [36]. Further, a small but growing body of research examining online information-seeking habits of medical professionals has found that Wikipedia is a popular resource for education or for finding answers to specific questions, primarily due to ease of access and use [37–40]. However, high utilization does not necessarily correspond to trust: these same samples tend to rank Wikipedia low on trustworthiness and credibility [37,39].

While Wikimedia sites seem poised to become the next frontier of health information dissemination efforts in medicine [41], comparable work in the mental health space is scant. Organizations and groups like Helping Give Away Psychological Science (HGAPS) and the Society for Clinical Psychology and the Society for Clinical Child and Adolescent Psychology (Divisions 12 and 53 of the American Psychological Association, respectively) recently began seeking to expand the availability of evidence-based psychological resources on open-source platforms like Wikipedia [42,43]. However, we know little about how well these efforts actually reach mental health clinicians, nor do we know how mental health clinicians use and evaluate Wikipedia and Wikiversity. An important first step in learning how to optimize the impact of dissemination efforts through Wikimedia and other internet platforms is to understand clinicians’ practices and perspectives. This study collected data from practicing mental health clinicians about where and how they seek information about psychological science on the Internet, and about ways that their confidence in the quality of online resources could be improved.

## Results

### Participant Demographics

Of the 133 survey respondents, 13 were excluded for incomplete responses (<50% complete). The final sample (*N* = 120) was 82.5% female, 13.3% male, and 1.7% identified as non-binary or third gender. Most respondents identified as White (85.6%); 9.3% identified as Black or African American, and 5.9% as Asian. No participants identified as Hispanic or Latinx. Respondents were 41.31 years old on average (*SD* = 13.81) and 53.3% had received a doctorate degree, 42.5% a Masters, and 0.8% a Bachelors (1.7% preferred not to disclose). Most reported their professional discipline as clinical psychology (60.0%), with additional representation from social work (25.8%), counseling (6.7%), school psychology (1.7%), and psychiatry (0.8%); 2.5% preferred to self-describe their discipline. Participants’ primary work settings were private practice (38.3%), hospital/academic medical centers (30.8%), schools/universities (8.3%), and community mental health centers (8.3%); 9.2% endorsed “other” work setting (e.g., primary care, prison settings).

### Information Seeking Patterns

Table 1 shows clinicians’ information-seeking patterns. Colleagues were the most common information source clinicians reported using when faced with mental health-related questions, with 80.0% often or almost always asking colleagues. Other commonly reported sources that were used often or almost always included Google (N= 68; 56.7%), academic journals (N = 51; 42.5%), Google Scholar (N = 47; 39.1%), textbooks (N = 44; 36.6%), and professional listservs (N = 43; 35.9%). Under a quarter of respondents (20.8*%*) reported often or almost always looking to PubMed and 10.8% to WebMD. Less than 1% of respondents reported often or almost always using Wikipedia or Wikiversity. Twitter was the least commonly endorsed source of information, with zero respondents often or almost always using it to seek mental health related information.

**Table 1.**
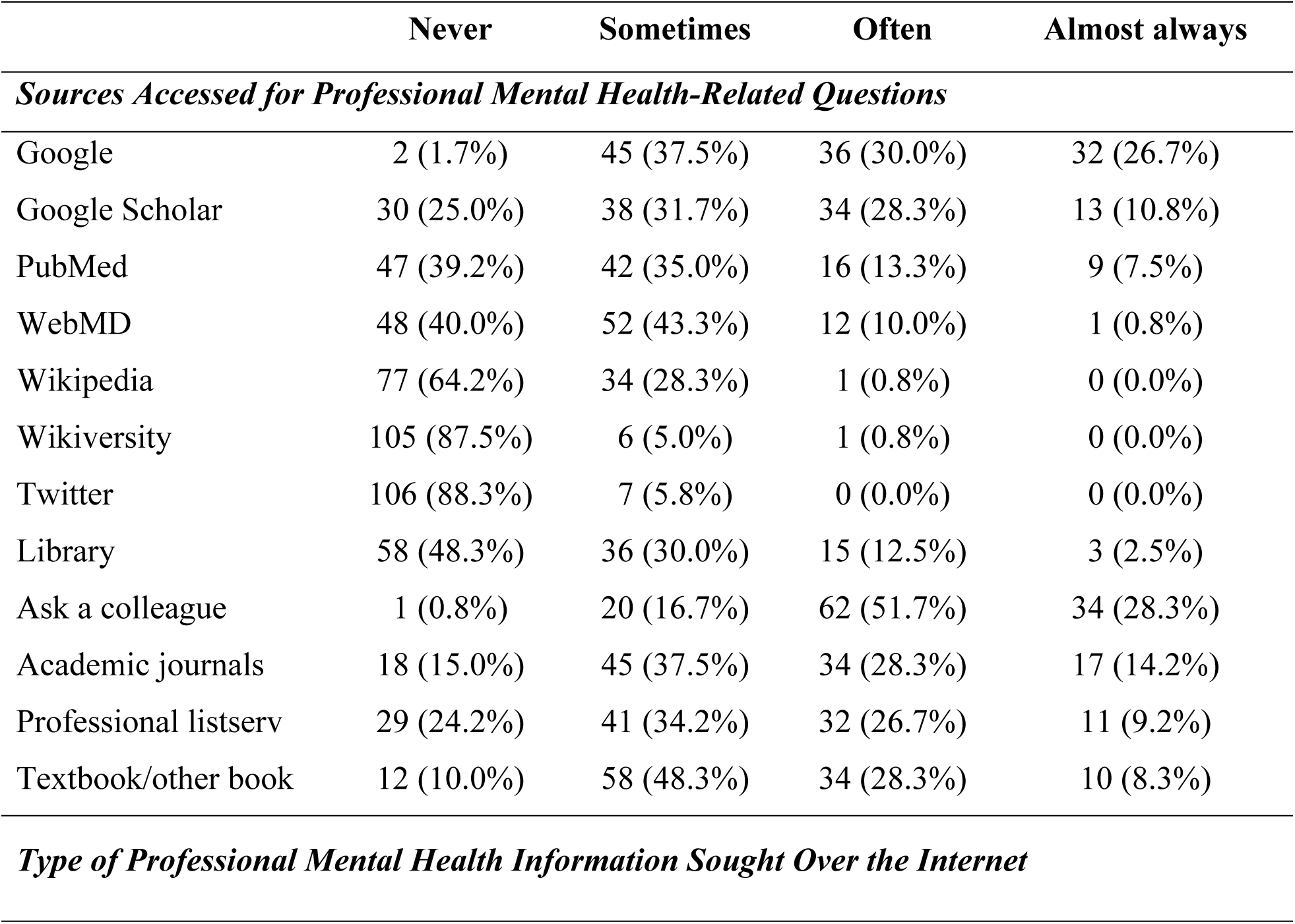
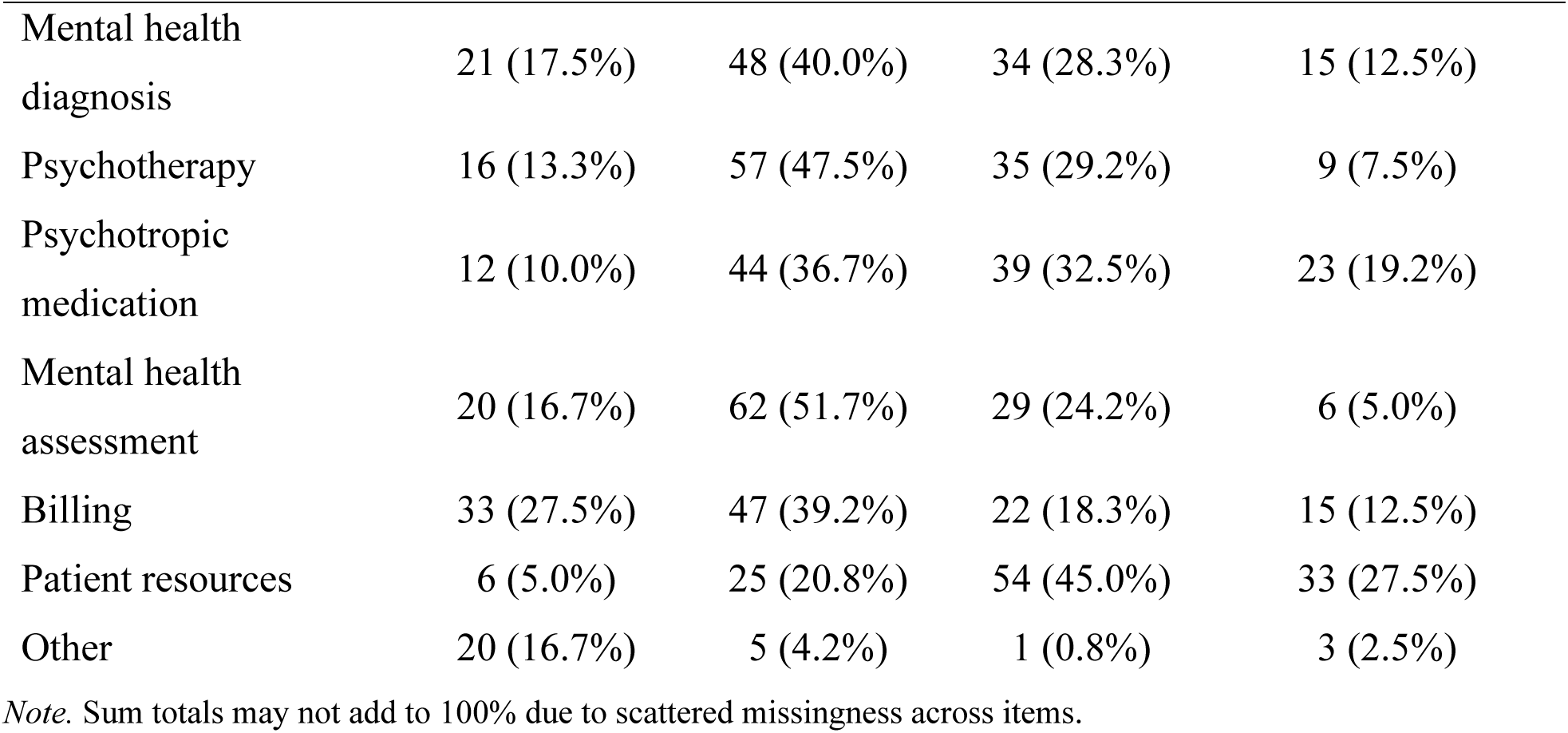
Information Seeking Patterns Reported by Clinician Sample (*N* = 120)

Most clinicians reported often or almost always (N = 87; 72.5%) using the internet for questions regarding resources to give to their patients. Other uses included searching for psychotropic medication-related questions (51.7% often or almost always), mental health diagnosis questions (40.8%), psychotherapy questions (36.7%), and billing (30.8%). Questions about mental health assessment had the lowest internet use rates (29.2% often or almost always searching) besides the write in “other” option. Examples of “other” options provided included “to investigate what professionals are available in specific areas of practice, to look up trainings I’m interested in,” “intervention resources and activities,” “resources to use in session like worksheets, activities, etc.,” and “specialized referral options.”

### Familiarity and Confidence Levels in Wiki Platforms

Table 2 reports respondent-reported familiarity and confidence in open-source platforms. Respondents had more familiarity with Wikipedia (100% of participants reported being aware of the resource; 73.4% very or extremely familiar) than Wikiversity (75.0% reported being aware of the resource; 2.5% very or extremely familiar); however, overall confidence in each platform was moderate to low (Wikipedia *M* = 3.78, *SD* = 3.48; Wikiversity *M* = 2.95, *SD* = 3.21).

**Table 2.** Familiarity with and Confidence Levels in Open-Source Platforms.

| Source | Familiarity, N (%) |  |  |  | Confidence, *M* (*SD*) |
| --- | --- | --- | --- | --- | --- |
|  | Not at all | Somewhat | Very | Extremely |  |
| Wikipedia | 8 (6.7%) | 20 (16.7%) | 47 (39.2%) | 41 (34.2%) | 3.78 (3.48) |
| Wikiversity | 102 (85.0%) | 12 (10.0%) | 1 (0.8%) | 2 (1.7%) | 2.95 (3.21) |

*Note.* Sum totals may not add to 100% due to scattered missingness across items. Confidence was measured on a 7-point Likert scale (1 = *least confidence*, 7 = *most confident*).

### Wiki Engagement for Mental Health Related Questions

Despite most clinicians reporting at least some familiarity with Wikipedia, many respondents (62.5%) reported never engaging with Wikipedia for mental health-related questions (see Table 3). For those that did engage with Wikipedia, questions about psychotropic medication were most searched for on Wikipedia, followed by questions related to diagnosis, psychotherapy, mental health assessment, billing, and patient resources. Most participants (70.0%) reported never directly searching Wikiversity for mental health-related questions. For the minority of respondents who reported directly engaging with Wikiversity (3.3%), questions about psychotherapy and patient resources were most common, followed by those related to mental health assessment, diagnosis, and psychotropic medication.

**Table 3.** How Clinicians Report Using Wikipedia and Wikiversity *(N* = 118)

|  | Never | Sometimes <sup>a</sup> | Often | Almost Always | N/A, never heard of this resource |
| --- | --- | --- | --- | --- | --- |
| ***Wikipedia*** |  |  |  |  |  |
| Directly search | 75 (62.5%) | 41 (34.2%) | 2 (1.7%) | 0 (0.0%) | 0 (0.0%) |
| ***Topics Searched<sup>b</sup>*** |  |  |  |  |  |
| Diagnosis | 22 (51.2%) | 19 (44.2%) | 2 (4.7%) | 0 (0.0%) | -- |
| Psychotherapy | 26 (60.5%) | 16 (37.2%) | 0 (0.0%) | 1 (2.3%) | -- |
| Psychotropic Medication | 17 (39.5%) | 20 (46.5%) | 5 (11.6%) | 1 (2.3%) | -- |
| Mental health assessment | 30 (69.8%) | 12 (27.9%) | 1 (2.3%) | 0 (0.0%) | -- |
| Billing | 33 (76.7%) | 10 (23.3%) | 0 (0.0%) | 0 (0.0%) | -- |
| Patient resources | 34 (79.1%) | 7 (16.3%) | 0 (0.0%) | 2 (4.7%) | -- |
| ***Wikiversity*** |  |  |  |  |  |
| Directly search | 84 (70.0%) | 4 (3.3%) | 0 (0.0%) | 0 (0.0%) | 30 (25.0%) |
| ***Topics Searched<sup>b</sup>*** |  |  |  |  |  |
| Diagnosis | 2 (50.0%) | 2 (50.0%) | 0 (0.0%) | 0 (0.0%) | -- |
| Psychotherapy | 0 (0.0%) | 4 (100.0%) | 0 (0.0%) | 0 (0.0%) | -- |
| Psychotropic Medication | 2 (50.0%) | 2 (50.0%) | 0 (0.0%) | 0 (0.0%) | -- |
| Mental health assessment | 1 (25.0%) | 2 (50.0%) | 1 (25.0%) | 0 (0.0%) | -- |
| Billing | 3 (75.0%) | 1 (25.0%) | 0 (0.0%) | 0 (0.0%) | -- |
| Patient resources | 1 (25.0%) | 3 (75.0%) | 0 (0.0%) | 0 (0.0%) | -- |

*Note.* Responses based on sample size of 118; two respondents left these items blank.
<sup>a</sup>Answers for “Rarely” and “Sometimes” were combined under “Sometimes” for the “Directly search” rows for parsimony in presentation.
<sup>b</sup>Questions regarding specific topics searched on each site are presented only for respondents who reported searching the site at least sometimes (Wikipedia $N = 43$ ; Wikiversity $N = 4$ ).

### Support for Verification Steps to Increase Confidence

No potential verification step was universally endorsed. However, many clinicians endorsed seeing Wiki pages linked on reputable mental health sites (N = 90, 76.3%) or a check mark indicating verified content (N = 88, 74.6%) as steps that would increase confidence. Seeing author names (N = 82, 69.5%) and the provision of some sort of quality rating based on articles cited received support from over half the sample. Eight respondents (6.8%) opted to describe an additional verification step; these included: knowing it is vetted by experts, including background information on the authors, having trusted colleagues discuss the resource, or if the information was cited for further investigation/review. One write-in response stated that all would be helpful; the other three declined to provide an additional verification step, citing unfamiliarity with Wiki platforms. Six (5.1%) respondents reported that no verification steps would increase their confidence in the content presented on these platforms. If indicated verification steps were implemented, 21.7% (N = 26) said they would be extremely or very likely to then turn to Wiki sources as reputable information sources and 68.3% (N = 82) reported they would be likely or somewhat likely to use them; 8.3% (N = 10) said that they would not at all be likely to use these sources even if verification steps were implemented.

## Discussion

To our knowledge, this is the first study to ask mental health clinicians directly about how and when they use Wikimedia platforms for mental health-related information. Our results showed that, despite increasingly being used to disseminate psychological information, Wiki sites are not seen as reputable resources by mental health clinicians. Colleagues were reported as the number one source participants turned to when faced with mental health-related questions.

General internet resources (e.g., Google searches) were the second most utilized resource. Interestingly, this latter finding may suggest knowledge or perception gaps, since respondents may or may not be aware that a Google search (the most commonly endorsed strategy) includes Wikipedia pages in its results both explicitly (by boosting the link to the first screen of results) and via scraping and embedding the content in large language models such as ChatGPT and also applications such as the “knowledge panels” that provide crisply formatted summaries [44].

Respondents reported higher confidence in, and greater familiarity with, Wikipedia than Wikiversity. However, most respondents reported never directly searching either for mental health-related information. With respect to information verification steps that might increase participants’ likelihood of using these platforms, most respondents said certain steps could increase their level of confidence in the information presented on Wikipedia and Wikiversity. Respondents also indicated that they would be more likely to use Wikipedia and Wikiversity if the verification steps were implemented, although overall rates of predicted utilization remained low. These findings align with previous research with other disciplines that professionals are wary of using open-source platforms, due in part to the perception that the information is not reliable or lacks proper oversight by field experts [21,28,29]. This remains true even among the increasing number of medical professionals who report high Wikipedia use [37,39,40]. It is worth noting that confidence ratings referred to the entirety of Wikipedia or Wikiversity; it is possible credibility perceptions would differ if respondents were asked to rate specific content pages.

This study sets a foundation for future research aimed at determining if Wiki platforms can meaningfully become an engine for dissemination of mental health resources to mental health professionals as it appears to be becoming for other medical professionals. Further studies building on these results should test whether addressing barriers to trusting and using Wiki platforms (e.g., implementing verification steps to increase confidence in the content presented) would lead to increased use. Future studies should also examine clinicians’ rationale for their perceived confidence in information from different sources. Understanding why some sites are trusted and others are not will inform future dissemination efforts that aim to leverage open-source platforms.

This study has several strengths. Most notably, this is the first study to ask mental health clinicians directly about how and when they use online platforms and can inform efforts to disseminate psychological science quickly and efficiently to practicing, community clinicians. Further, we had wide variety in respondent degree level, professional discipline, and primary work settings, allowing for this research to capture opinions from a diverse range of professional experience.

Several study limitations are important to note. Most notably, respondents represented a convenience sample of practicing clinicians recruited via online listservs. As such, participants may be more interested in the topic of dissemination than the broader clinician population. This suggests that, if anything, our results may overestimate the confidence and frequency with which clinicians use Wiki platforms for mental health-related questions. Our sample also had a larger proportion of doctoral-level providers compared to the primarily master’s level mental health workforce [45]. As such, although this study provides important preliminary data on skepticism of open-source platforms among practicing clinicians, the findings are not generalizable to the population of clinicians at large; future work with a more diverse representation of clinicians is needed. However, given the low confidence in Wiki platforms endorsed by this sample, it is unlikely that sample representativeness undermines our central finding that the goal of using open-source platforms like Wikipedia to disseminate new knowledge in the field of mental health faces a steep reputational obstacle. Finally, we relied on participants’ report of their resource searching habits; some participants may have been motivated to downplay their use of these sites – particularly in the context of a survey conducted by other mental health professionals. Discrepancies between self-report of online behavior and objective tracking of online behavior are common and often large (see [46] for examples). Future studies should combine assessment strategies for indexing Wikimedia use.

Overall, this study indicates that mental health clinicians are not regularly using Wiki sites for mental health-related questions, even though they are accessible, regularly updated, and increasingly reliable. Findings suggest that implementing verification steps on Wiki pages could increase clinicians’ confidence in the information provided and their likelihood in using these sites as resources. Increasing awareness about the use of Wiki sites and open science platforms in other disciplines, including health-related topics such as COVID-19, and independent evaluations of accuracy, all may be helpful in changing perceptions and acceptability. However, it remains unclear whether verification steps would lead to more frequent use or how such a system would be implemented through existing platforms. Overall, these results will help guide future dissemination efforts to increase the availability and utilization of evidence-based psychological science online for clinicians.

## Materials and Methods

### Participants

Survey invitations were sent via several listservs for mental healthcare providers, including the Association for Behavioral and Cognitive Therapies (ABCT), the Anxiety & Depression Association of America (ADAA), the North Carolina Psychological Association, the Area Health Education Centers Program in North Carolina, a local social work listserv in Philadelphia, PA, as well as word of mouth. Follow-up emails were sent to listservs to remind recipients about the one-time, electronic survey. The initial sample size of completed surveys included *N*=133 practicing mental health clinicians. Of the 133 survey respondents, 13 were excluded for incomplete responses (<50% complete), resulting in a final sample size of *N* = 120.

### Procedure

We developed a survey to descriptively assess respondent awareness of and confidence in Wikipedia and Wikiversity, among other online resources. We asked specifically about these platforms because they have been the focus of distinct mental health information dissemination initiatives [47] and have been studied as a metric of the reach and impact of psychological research [48]. Wikipedia is visited over 20 billion times per month [8]. Wikiversity is less visited, but more geared towards technical information, and it has been a priority for disseminating information to mental health professionals and trainees. Prior to implementation, the survey was reviewed by four experts in psychological resources and iteratively revised before finalizing to ensure broad representation of available information sources and information-seeking habits, as well as clarity of the questions and response options.

The final survey consisted of 18 multiple choice questions about information seeking behavior and confidence in online resources, followed by demographic and clinical background questions. The survey was administered via Qualtrics, an online data management system, and took approximately 10 to 15 minutes to complete. All respondents provided informed consent prior to participation. Participants were given the option of receiving a $5 electronic gift card as compensation. Procedures were approved by the Institutional Review Board at the University of Pennsylvania.

### Survey Items

#### Information Seeking Patterns

Participants first were presented with a list of twelve information sources and asked to indicate the frequency (“Never,” “Sometimes, “Often,” or “Always”) with which they look to each for answers/resources when faced with a mental health-related question at work.

Participants were then asked to rate how often they typically used the Internet to search for answers to specific questions about mental health care (e.g., diagnosis, assessment), on the same scale.

#### Familiarity and Confidence Levels in Wiki Platforms

Following report of general online information seeking patterns, participants were asked to rate their level of familiarity with Wikiversity and Wikipedia on a 4-point Likert scale from “Not at all familiar” to “Extremely familiar.” Participants also were asked to rate how confident they felt about the mental health information that is available from each of those sources on a 7- point Likert scale (1 = not at all confident, 7 = very confident).

#### Wiki Engagement for Mental Health Related Questions

We then asked how often participants search for answers to questions about mental health on Wikiversity and Wikipedia. Respondents who indicated that they did search for mental health content on these sites were provided with a list of seven mental health topics (with one option for write in listed as “Other”) and asked how often they searched each site for each mental health topic. Answers were given on a 5-point Likert scale as “Never,” “Rarely,” “Sometimes,” “Often,” or “Always.”

#### Support for Verification Steps to Increase Confidence

Finally, participants were asked what might increase their level of confidence in the information presented on Wikipedia and Wikiversity (e.g., a check mark for verified content, quality rating based on articles cited). Clinicians could also write in alternative options or specify that “nothing would increase my confidence.” Participants were then asked how likely they would be to use platforms like Wikipedia or Wikiversity for information-gathering if the verification steps they had selected were implemented. Answers were given on a 5-point Likert scale as “Not at all likely,” “Somewhat likely,” “Likely,” “Very Likely,” or “Extremely Likely.”

#### Participant Demographics

Participants reported on demographic (age, gender, race, ethnicity) and clinical and professional background characteristics (e.g., education background, professional discipline, primary work setting).

### Data Analysis Plan

We produced descriptive statistics for Likert-based questions to address our study aims. Answers indicating “Rarely” or “Sometimes” were combined for the two questions regarding how often clinicians engage with Wikipedia and Wikiversity to search for answers to questions about mental health.

## Data Availability

Data are available upon request from first author SY.

